# A Benchmark Dose Analysis for Maternal Pregnancy Urine-Fluoride and IQ in Children

**DOI:** 10.1101/2020.10.31.20221374

**Authors:** Philippe Grandjean, Howard Hu, Christine Till, Rivka Green, Morteza Bashash, David Flora, Martha Maria Tellez-Rojo, Peter Song, Bruce Lanphear, Esben Budtz-Jørgensen

**Author notes:** Address correspondence to Philippe Grandjean, Environmental Medicine, University of Southern Denmark, J.B.Winslowsvej 17A, 5000 Odense C, Denmark.

## Abstract

As a safe exposure level for fluoride in pregnancy has not been established, we used data from two prospective studies for benchmark dose modeling. We included mother-child pairs from the Early Life Exposures in Mexico to Environmental Toxicants (ELEMENT) cohort in Mexico and the Maternal-Infant Research on Environmental Chemicals (MIREC) cohort in Canada. Children were assessed for IQ at age 4 (n=211) and between 6 and 12 years (n=287) in the ELEMENT cohort and between ages 3 and 4 years (n=512) in the MIREC cohort. We calculated covariate-adjusted regression coefficients and their standard errors to explore the concentration-effect function for maternal urinary fluoride with children’s IQ, including possible sex-dependence. Assuming a benchmark response of 1 IQ point, we derived benchmark concentrations (BMCs) of maternal urinary fluoride and benchmark concentration levels (BMCLs). No deviation from linearity was detected from the results of the two studies. Using a linear slope, the BMC for maternal urinary fluoride associated with a 1-point decrease in IQ scores of preschool-aged boys and girls was 0.29 mg/L (BMCL, 0.18 mg/L). The BMC was 0.30 mg/L (BMCL, 0.19 mg/L) when pooling the IQ scores from the older ELEMENT children and the MIREC cohort. Boys showed slightly lower BMC values compared with girls. Relying on two prospective studies, maternal urine-fluoride exposure at levels commonly occurring in the general population, the joint data showed BMCL results about 0.2 mg/L. These results can be used to guide decisions on preventing excess fluoride exposure in vulnerable populations.

## 1. INTRODUCTION

In 2006, the U.S. National Research Council (NRC) evaluated the US Environmental Protection Agency’s (EPA) fluoride standards and concluded that fluoride can adversely affect the brain (National Research Council 2006). The EPA’s Maximum Contaminant Level Goal (MCLG) of 4.0 mg/L was set to protect against crippling skeletal fluorosis. In 2020, a meta-analysis of epidemiological studies was conducted by the National Toxicology Program (National Toxicology Program 2020), but it did not distinguish between prenatal and postnatal exposure and did not consider the full information from the most recent prospective evidence. A substantial number of cross-sectional studies, mostly in communities with chronic fluoride exposure, have shown cognitive deficits in children growing up in areas with elevated fluoride concentrations in drinking water (Choi et al. 2012; Choi et al. 2015; Tang et al. 2008; Duan et al. 2018), and this evidence is supported by experimental toxicology studies (Mullenix et al. 1995; Bartos et al. 2018; National Toxicology Program (NTP) 2016).

Fluoride is found in many minerals and in soil (National Research Council 2006), and thus also in groundwater. Since the mid-1940s, fluoride has been added to many community water supplies with the aim of preventing tooth decay (U.S. Environmental Protection Agency (U.S. EPA) 1985). Community water fluoridation is applied in the U.S. and Canada and several other countries, whereas other countries, like Mexico, have chosen to add fluoride to table salt. Community water fluoridation accounts for about 40-70 percent of daily fluoride intake in adolescents and adults living in fluoridated communities (U.S. Environmental Protection Agency (U.S. EPA) 2010), with fluoride concentration of drinking water roughly equaling the fluoride concentration in urine (National Research Council 2006). In a cohort of pregnant women in Canada, the fluoride concentration in tap water showed a strong correlation with urinary fluoride excretion (r =.51)(Till et al. 2018). In addition to fluoridated water, some types of tea, such as black tea, are an important source of exposure (Waugh et al. 2017; Rodríguez et al. 2020; Krishnankutty et al. 2020).

Fluoride is readily distributed throughout the body, with bones and teeth as storage depots. During pregnancy, fluoride also crosses the placenta and reaches the fetus (World Health Organization 2006; National Research Council 2006). As fluoride is rapidly eliminated via urine, the volume-adjusted urine-fluoride (U-F) concentration mainly represents recent absorption (World Health Organization 2006; Ekstrand and Ehrnebo 1983). Pregnant women may show lower U-F concentrations than non-pregnant controls, perhaps due to fetal uptake and storage in hard tissues (Opydo-Symaczek and Borysewicz-Lewicka 2005); the U-F excretion tends to increase from the first to the third trimester (Till et al. 2020), a tendency that needs to be taken into account.

For the purpose of identifying safe exposure levels, regulatory agencies routinely use benchmark dose (BMD) calculations (EFSA Scientific Committee (EFSA) 2009; U.S. Environmental Protection Agency (U.S. EPA) 2012). As long recognized (National Research Council (NRC) 1989), fluoride is not an essential nutrient, and dose-dependent toxicity can therefore be considered monotonic. As with lead (Budtz-Jorgensen et al. 2013), BMD results can be generated from regression coefficients and their standard errors for the association between maternal U-F concentrations and the child’s IQ score (Grandjean 2019). The BMD is the exposure dose leading to a specific change (denoted BMR) in the response (in this case, an IQ loss), compared with unexposed children. A decrease of 1 IQ point is an appropriate BMR, as specified by the European Food Safety Authority (EFSA) and also recognized by the U.S. EPA (European Food Safety Authority 2010; Gould 2009; Reuben et al. 2017; Budtz-Jorgensen et al. 2013). We have chosen to use the data from two recently conducted, robust prospective birth cohort studies (Bashash et al. 2017; R. Green et al. 2019) to calculate the concentration of urine-fluoride (U-F) associated with a 1-point decrement in Full Scale IQ (FSIQ), which is considered a highly significant endpoint from a public health (Lanphear 2015) and an economic standpoint (Gould 2009).

## 2. METHODOLOGY

### 2.1. Study Cohorts

In the Early Life Exposures in Mexico to Environmental Toxicants (ELEMENT) project, mother– child pairs were successively enrolled in longitudinal birth cohort studies from the same three hospitals in Mexico City which serve low to moderate income populations. A full description of the cohorts and associated methods is provided in a recent “Cohort Profile” paper (Perng et al. 2019). Eligible mothers for whom urinary samples were collected during pregnancy were recruited between 1997 and 1999 in cohort 2A (n=237) and between 2001 and 2003 in cohort 3 (n=670). Participants were included if they had at least one biobanked urine sample available for fluoride analysis, a urinary creatinine concentration, and if their child underwent cognitive testing at age 4 years (n=287) and/or between ages 6 and 12 years (n=211).

In the Maternal-Infant Research on Environmental Chemicals (MIREC) program, 2,001 pregnant women were recruited between 2008 and 2011 from ten cities across Canada, seven of which have water fluoridation, while three do not. Women were recruited from prenatal clinics if they were at least 18 years old, less than 14 weeks gestation, and spoke English or French. Exclusion criteria included fetal abnormalities, medical complications, and illicit drug use during pregnancy; further details have been previously described (Arbuckle et al. 2013). A subset of children (n = 601) in the MIREC Study was evaluated for the developmental phase of the study (MIREC-Child Development Plus) at three-to-four years of age from six of the ten cities included in the original cohort, half of which were fluoridated. Of the 601 children who completed the neurodevelopmental testing in entirety, 512 (85.2%) mother-child pairs had all three U-F samples and complete covariate data to be included in the analyses; 75 (12.5%) women were missing one or more trimester U-F samples, and 14 women (2.3%) were missing one or more covariates.

### 2.2. Exposure Assessment

All urine samples from the two studies were analyzed by the same laboratory at the Indiana University School of Dentistry using a modification of the hexamethyldisiloxane (Sigma Chemical Co., USA) microdiffusion method with the ion-selective electrode (Martinez-Mier et al. 2011).

In the ELEMENT study, spot (second morning void) urine samples were collected during the first trimester (mean ± SD: 13.6 ± 2.1 weeks for cohort 3 and 13.7±3.5 weeks for cohort 2A), second trimester (25.1 ± 2.3 weeks for cohort 3 and 24.4 ± 2.9 weeks for cohort 2A), and third trimester (33.9 ± 2.2 weeks for cohort 3 and 35.0 ± 1.8 weeks for cohort 2A). The samples were collected into fluoride-free containers and immediately frozen at the field site and shipped and stored at −20°C at the Harvard School of Public Health (HSPH), and then at −80°C at the University of Michigan School of Public Health (UMSPH). To account for variations in urinary dilution at time of measurement, we adjusted maternal U-F concentration for urinary creatinine, as described in our prior work (Thomas et al. 2016). An average of all available creatinine-adjusted U-F concentrations during pregnancy (maximum 3 samples and minimum one sample) was computed and used as the exposure measure.

In the MIREC study, urine spot samples were collected at each trimester, i.e., first trimester at 11.6 ± 1.6 (mean ± SD) weeks of gestation, second trimester at 19.1 ± 2.4 weeks, and third trimester at 33.1 ± 1.5 weeks. Maternal U-F concentrations at each trimester were adjusted for specific gravity, as described in our previous work (Till et al. 2020). Creatinine-adjusted U-F concentrations were also derived for each woman, but U-F concentrations adjusted for specific-gravity were preferred because more data were available and adjustment for creatinine did not substantially alter the results (R. Green et al. 2019). For each woman, the average maternal U-F concentration was derived only if a valid U-F value was available for each trimester.

### 2.3. Assessment of Intelligence

The ELEMENT study (Bashash et al. 2017) used the McCarthy Scales of Children’s Abilities (MSCA) Spanish version to measure cognitive abilities at age 4 years and derive a General Cognitive Index (GCI) as a standardized composite score. The MSCA was administered by trained psychometrists or psychologists who were supervised by an experienced clinical child psychologist. The GCI shows concurrent validity with intelligence tests, including the Stanford-Binet IQ (r=.81) and FSIQ (r=.71) from the Wechsler Preschool and Primary Scale of Intelligence (WPPSI) (Kaplan and Sacuzzo 2010). For children age 6-12 years, a Spanish-version of the Wechsler Abbreviated Scale of Intelligence (WASI) was administered to derive FSIQ as a measure of global intellectual functioning. In the MIREC study, children’s intellectual abilities (R. Green et al. 2019) were assessed at ages 3 to 4 years using the FSIQ from the Wechsler Preschool and Primary Scale of Intelligence, Third Edition (WPPSI-III). A trained research assistant who was supervised by a psychologist administered the WPPSI-III in either English or French. In both studies, examiners were blinded to the children’s fluoride exposure. All raw scores were standardized for age and sex.

### 2.4. Covariate Adjustment

For the ELEMENT study, data were collected from each subject by questionnaire on relevant parameters, gestational age was estimated by registered nurses, and maternal IQ was estimated using subtests of the Wechsler scale standardized for Mexican adults. The quality of the children’s individual home environments was assessed using an age-appropriate version of the Home Observation for the Measurement of the Environment (HOME) score (Caldwell and Bradley 1984). Covariates included gestational age, birth weight, sex, HOME, age at outcome measurement and the following maternal characteristics: parity, smoking history, marital status, age at delivery, IQ, education, and sub-cohort.

The MIREC study selected similar covariates from a set of established predictors of fluoride metabolism and cognitive development, including maternal age, sex, city of residence, HOME score, maternal education, and maternal race/ethnicity. Covariates included in the original studies (Bashash et al. 2017; R. Green et al. 2019) were retained in the statistical calculations in the present study. Due to a growing body of epidemiologic studies showing sex-specific effects associated with neurotoxic exposures, including fluoride (R. Green et al. 2019; R. Green et al. 2020), interactions between sex and U-F exposure were considered.

### 2.5 Benchmark Concentration (BMC) Calculations

The BMC is the U-F concentration that reduces the outcome by a prespecified level (known as the benchmark response, BMR) compared to an unexposed control with the same covariate profile (20, 21). We based the benchmark calculations on regression models in the following form:

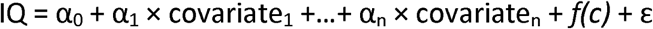

where c is the fluoride concentration and *f* is the concentration-response function. To assess the linearity of the concentration-response relationship, several models were estimated. In addition to the standard linear model, where *f(c)* = βc, we estimated a squared effect, where *f(c)* = βc^2^, and two piecewise-linear models with breakpoints at 0.5 and 0.75 mg/L. Piecewise-linear models are useful in benchmark calculations because the slope of the concentration-response function is allowed to change at the breakpoint and therefore in such models benchmark calculations are not sensitive to exposure effects occurring only at high concentration levels. Furthermore, to allow for the possibility of different exposure effects in boys and girls, each concentration-response model was also fitted with the inclusion of an interaction with sex. Models were fitted independently in the two cohorts yielding analyses that were similar to those presented in the original publications (Bashash et al. 2017; R. Green et al. 2019) based on the original raw data and with the covariate adjustments as originally justified. From the regression results, we first calculated BMC results for each cohort and we then derived joint BMCs by combing regression coefficients from the two cohorts.

A smaller BMR will result in lower BMC and benchmark concentration level (BMCL) results. In this study, the outcome variable is child IQ and we selected a BMR of 1 IQ point to derive the BMC. In our regression model, the IQ difference between unexposed subjects and subjects at the BMC is given by f(0)− f(BMC), and therefore the BMC satisfies the equation f(0)− f(BMC)=BMR. We use concentration-response functions with f(0)=0, and therefore the BMC is given by

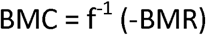

In a regression model with a linear concentration-response function [f(c) = βc], we get BMC = −BMR/β. If the estimated concentration-response is increasing (indicating a beneficial effect), the BMC is not defined, and the BMC is then indicated with a ∞.

The main result of the BMC analysis is the BMCL, which is defined as a lower one-sided 95% confidence limit of the BMC. In the linear model

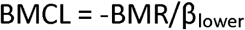

where β_lower_ is the one-sided lower 95% confidence limit for β (Budtz-Jorgensen et al. 2013). In the other models considered, we calculated the BMCL by first finding a lower confidence limit for *f(c)* and then finding the concentration (*c*) where confidence limit is equal to -BMR.

Finally, we derived two sets of joint benchmark concentrations: MIREC results (FSIQ score) were combined with ELEMENT outcomes using either GCI or FSIQ scores. Joint benchmark concentration results were obtained under the hypothesis that the concentration-response functions were identical in the two studies. Under this hypothesis, the concentration-response function [*f(c)*] was estimated by combing the regression coefficients describing *f(c)*. Again, using the linear model as an example, we estimated the joint regression coefficient by weighing together cohort specific coefficients. Here we used optimal weights proportional to the inverse of the squared standard error. In a Wald test, we tested whether the exposure effects in the two cohorts were equal. We calculated sex-dependent BMC results from regression models including interaction terms between sex and *f(c)*. The fit of the regression models was compared by twice the negative log-likelihood [−2 log*L*], where a lower value indicates a better fit. As the linear model is nested in the piecewise linear model, the fit of these two models could be compared using likelihood ratio testing. Thus, we calculated the p-value for the hypothesis that the concentration-response is linear in a test where the alternative was the piecewise linear model. Here a low p-value indicates that the linear model has a poorer fit.

## 3. RESULTS

Table 1 shows the regression coefficients obtained from the two outcomes (GCI and IQ score) in the ELEMENT study and the IQ score in the MIREC study. While the MIREC Study did not show a statistically significant association with U-F exposure for boys and girls combined, the joint regression coefficients for the MIREC and ELEMENT studies combined are significant, and the deviations between the studies are not. These results are consistent across the linear and squared concentration scales.

**Table 1.** Regression coefficients adjusted for confounders for the change in the outcome at an increase by 1 mg/L in fluoride concentration for IQ in the MIREC study, GCI (upper rows) and IQ (lower rows) in the ELEMENT study, and a joint calculation. The column to the right shows the p-value for a hypothesis of identical regressions in the two studies. Two dose-response models are used, a linear and one with the squared exposure variable.

|  | MIREC |  | ELEMENT |  | Joint |  |  |
| --- | --- | --- | --- | --- | --- | --- | --- |
| model | beta | p | Beta | p | beta | P | P<sub>diff</sub> |
|  | FSIQ (n=512) |  | GCI (n=287) |  |  |  |  |
| Linear | -2.07 | 0.211 | -6.29 | 0.007 | -3.49 | 0.01 | 0.139 |
| Squared | -0.963 | 0.232 | -2.68 | 0.02 | -1.53 | 0.021 | 0.221 |
|  | FSIQ (n=512) |  | IQ (n=211) |  |  |  |  |
| Linear | -2.07 | 0.211 | -5.00 | 0.01 | -3.30 | 0.009 | 0.252 |
| Squared | -0.963 | 0.232 | -2.65 | 0.002 | -1.75 | 0.003 | 0.152 |

Table 2 shows the BMC results obtained from the regression coefficients for both sexes, and separately by sex. The BMC and BMCL are presented for the MIREC study, the ELEMENT (GCI and IQ) study, and combined across the two studies. The joint model fits are approximately the same between the linear and squared models and for both IQ and GCI outcomes. For the joint linear models, the average BMCL in terms of U-F is approximately equal for the MIREC-ELEMENT IQ model (0.19 mg/L) and MIREC-ELEMENT GCI model (0.18 mg/L). Likewise, for the joint squared models, the average BMCL in terms of U-F is approximately equal for the MIREC-ELEMENT IQ model (0.61 mg/L) and MIREC-ELEMENT GCI model (0.62 mg/L).

**Table 2.** Benchmark dose results (mg/L urinary fluoride) for a BMR of 1 IQ point obtained from the MIREC study and the two cognitive assessments from the ELEMENT study as well as the joint results. Two dose-response models are used, a linear and one with the squared exposure variable. For both models, sex-specific and joint benchmark results are provided. The fit of the regression models were compared by twice the log-likelihood [−2 log*L*] where lower values indicate a better fit.

| Study |  | MIREC<br>(n=512) |  | ELEMENT IQ (n=211) |  | ELEMENT GCI<br>(n=287) |  | MIREC and ELEMENT IQ<br>(n=723) |  |  | MIREC and ELEMENT GCI<br>(n=799) |  |  |
| --- | --- | --- | --- | --- | --- | --- | --- | --- | --- | --- | --- | --- | --- |
| Model | Sex | BMD | BMDL | BMD | BMDL | BMD | BMDL | BMD | BMDL | -2logL | BMD | BMDL | -2logL |
| Linear | Both | 0.483 | 0.208 | 0.200 | 0.122 | 0.159 | 0.099 | 0.303 | 0.186 | 5541.9 | 0.286 | 0.175 | 6263.1 |
| Linear | Boys | 0.218 | 0.127 | 0.275 | 0.130 | 0.148 | 0.084 | 0.237 | 0.148 | 5536.3 | 0.192 | 0.125 | 6258.0 |
| Linear | Girls | ∞ | 0.515 | 0.160 | 0.091 | 0.169 | 0.087 | 0.691 | 0.218 | 5536.3 | 1.372 | 0.247 | 6258.0 |
| Squared | Both | 1.019 | 0.661 | 0.614 | 0.496 | 0.611 | 0.467 | 0.756 | 0.606 | 5540.5 | 0.809 | 0.619 | 6264.5 |
| Squared | Boys | 0.716 | 0.537 | 0.684 | 0.496 | 0.581 | 0.435 | 0.703 | 0.558 | 5535.5 | 0.666 | 0.533 | 6259.8 |
| Squared | Girls | ∞ | 1.158 | 0.576 | 0.449 | 0.642 | 0.434 | 0.921 | 0.609 | 5535.5 | 6.436 | 0.729 | 6259.8 |

Models allowing for sex-dependent effects showed better fit mainly due to the significant interaction terms in the MIREC cohort. Regarding sex-dependent effects, no apparent fluoride-associated IQ loss was seen in girls in the MIREC cohort. Although the BMCL is clearly higher in girls than boys (0.52 vs. 0.13 mg/L), the overall BMCL for both sexes in the MIREC cohort (0.21 mg/L) is closer to the one for boys than the one for girls.

Table 3 shows results of sensitivity analysis using piecewise linear functions, with one breakpoint at 0.75 mg/L and one at 0.5 mg/L. A piecewise linear model is more flexible than a linear model, but likelihood ratio testing showed that joint data piecewise linear models did not fit significantly better than the standard linear models. Thus, the hypothesis of a linear concentration-response relation could not be rejected: for the joint MIREC IQ and ELEMENT IQ data, p-values for likelihood testing were p=0.85 and p=0.90 when the linear model was tested against models using breakpoints of 0.5 and 0.75 mg/L, respectively. For the joint MIREC IQ and ELEMENT GCI, the corresponding p-values were p=0.18 and p=0.28.

**Table 3.** Benchmark concentration results (mg/L urinary fluoride) for a BMR of 1 IQ point obtained from the MIREC study and the two cognitive assessments from the ELEMENT study as well as the joint results. Two piecewise linear dose-response models (with urinary fluoride breakpoints at 0.5 and 0.75 mg/L) are used. For both models, sex-dependent and joint benchmark results are provided. The fit of the regression models was compared by twice the log-likelihood [−2 log*L*] where lower values indicate a better fit.

| Study | Sex | MIREC<br>(n=512) |  | ELEMENT<br>IQ (n=211) |  | ELEMENT<br>GCI (n=287) |  | MIREC and<br>ELEMENT IQ (n=723) |  |  | MIREC and<br>ELEMENT GCI (n=799) |  |  |
| --- | --- | --- | --- | --- | --- | --- | --- | --- | --- | --- | --- | --- | --- |
|  |  | BMD | BMDL | BMD | BMDL | BMD | BMDL | BMD | BMDL | -2logL | BMD | BMDL | -2logL |
| Breakpoint<br>0.5 | Both | 0.446 | 0.090 | 2.688 | 0.431 | 1.004 | 0.042 | 0.763 | 0.121 | 5538.5 | 0.616 | 0.099 | 6262.8 |
| Breakpoint<br>0.5 | Boys | 0.107 | 0.047 | 2.953 | 0.135 | 0.725 | 0.011 | 0.181 | 0.060 | 5532.6 | 0.108 | 0.048 | 6257.0 |
| Breakpoint<br>0.5 | Girls | ∞ | 0.121 | 2.363 | 0.024 | 1.144 | 0.046 | 1.433 | 0.135 | 5532.6 | 1.954 | 0.145 | 6257.0 |
| Breakpoint<br>0.75 | Both | 0.337 | 0.115 | 1.283 | 0.149 | 0.115 | 0.05 | 0.714 | 0.154 | 5539.4 | 0.240 | 0.108 | 6262.9 |
| Breakpoint<br>0.75 | Boys | 0.144 | 0.070 | 1.379 | 0.121 | 0.127 | 0.035 | 0.247 | 0.096 | 5533.6 | 0.142 | 0.071 | 6257.2 |
| Breakpoint<br>0.75 | Girls | 5.025 | 0.125 | 1.155 | 0.052 | 0.109 | 0.044 | 1.016 | 0.139 | 5533.6 | 0.398 | 0.108 | 6257.2 |

The shapes of linear, squared, and one piecewise concentration-response curves are shown in Figure 1. The figure shows that the squared model has weaker a slope at low doses, thus resulting in higher BMC results.

**Fig. 1.**
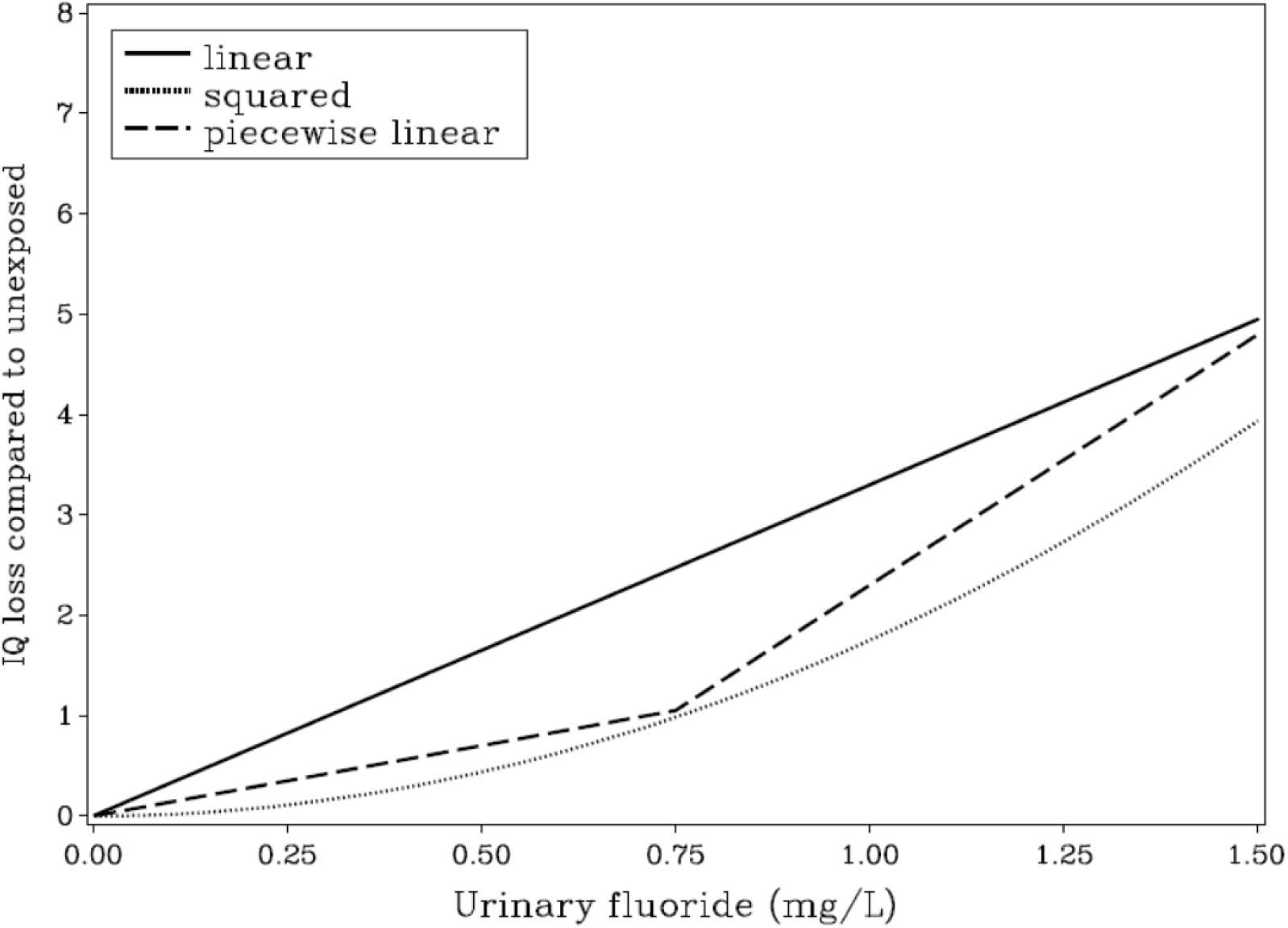
Association between maternal urinary-fluoride concentration in pregnancy and child IQ loss. Covariate-adjusted models shown for the linear (solid), squared (dotted), and piecewise (dashed) linear curve with breakpoint 0.75 mg/L. The BMD results and the curve fits are shown in Tables 2 and 3.

## 4. DISCUSSION

Experimental and cross-sectional epidemiology studies have provided evidence of fluoride neurotoxicity, especially when the exposure occurs during early brain development (Grandjean 2019). Evidence reviewed about 15 years ago suggested that sufficient information was available to warrant further consideration of the possible adverse effects of fluoride exposure with an emphasis on vulnerable populations (National Research Council 2006). Only now has thorough prospective epidemiology evidence become available on populations exposed to fluoridated water (about 0.7 mg/L) or comparable exposure from fluoridated salt. The present study is based on data from two birth cohort studies (R. Green et al. 2019; Bashash et al. 2017) that included detailed assessment of urinary fluoride concentrations during pregnancy. In these two studies, the mean U-F concentration was higher among pregnant women living in Mexico City (0.89 mg/L) as compared with the pregnant women living in fluoridated cities in Canada (0.69 mg/L). However, the range of U-F concentrations was similar, and the fluoride-associated cognitive deficits were similar, thus allowing joint calculations that provided better statistical power.

The MIREC study also estimated maternal fluoride intake in pregnancy from water and beverage consumption (e.g. tea, coffee) using questionnaires and measured fluoride concentration in municipal drinking water that services the mothers’ residences. The results showed similar IQ-loss associations, especially for non-verbal intelligence, for both fluoride intake and water fluoride concentrations (R. Green et al. 2019). Due to the brain’s continued vulnerability across early development (Grandjean 2013), early infancy may also be a vulnerable period of exposure for adverse effects from fluoride, especially among formula-fed infants who receive formula reconstituted with fluoridated water (Till et al. 2019). However, fluoride exposure in school-age children in the ELEMENT study showed a weaker and non-statistically significant adverse impact on the IQ in the cross-sectional analysis (Bashash et al. 2017). Taken together, these findings suggest that both prenatal and early childhood fluoride exposure may affect the development of IQ.

The magnitude of the fluoride-associated IQ losses are in accordance with findings in cross-sectional studies carried out in communities, where the children examined had likely been exposed to chronic water-fluoride concentrations throughout development (Choi et al. 2012). More recent studies have shown similar results (Yu et al. 2018; Wang et al. 2020), and benchmark dose calculations (Hirzy et al. 2016) relying on a large cross-sectional study (Xiang et al. 2003) showed results similar to the ones obtained in the current analysis. These findings provide additional evidence that fluoride is a developmental neurotoxicant (i.e., causing adverse effects on brain development in early life), similar to other toxic elements like lead, mercury and arsenic. However, blood concentrations of the latter elements associated with adverse effects are about 100-fold lower than serum-fluoride concentrations that correspond to the benchmark concentration (Grandjean 2019).

A few retrospective studies have been carried out in communities with fluoridation, though with imprecise exposure assessment that mostly relied on proxy variables, and without information on maternal fluoride exposure during pregnancy (Aggeborn and Ohman 2017; Broadbent et al. 2015). Further, in the ELEMENT cohort, elevated maternal U-F concentrations were associated with higher scores on inattention on the Conners’ Rating Scale, as indication of Attention-Deficit/Hyperactivity Disorder (ADHD) behaviors (Bashash et al. 2018). Other studies on attention outcomes found an association between water fluoridation and diagnosis of ADHD in Canada, although data on child U-F did not replicate this association (Riddell et al. 2019), consistent with the ELEMENT study of child U-F and IQ. Likewise, increased risk of ADHD was reported to be associated with water fluoridation at the state level in the U.S. (Malin and Till 2015), although inclusion of mean elevation at the residence as a covariate made the association non-significant (Perrott 2018).

Individual vulnerability may play a role, and the original MIREC study suggested that boys may be more vulnerable to prenatal fluoride neurotoxicity than girls (R. Green et al. 2019) suggesting a possible indication that sex-dependent endocrine disruption may play a role (Bergman et al. 2013), among other sex-differential possibilities. For example, genetic predisposition to fluoride neurotoxicity may exist (Zhang et al. 2015; Cui et al. 2018), but has so far not been verified. However, other predisposing factors, such as iodine deficiency (Malin et al. 2018) may well exist. For this reason, regulatory agencies use an uncertainty factor to derive safe exposure levels that are lower than the BMCL.

Both prospective studies adjusted for a substantial number of cofactors while others were found to be unimportant. Prenatal and early postnatal lead exposure did not influence the fluoride-associated IQ deficits. Other neurotoxicants or risk factors, such as arsenic and lead exposure, did not appreciably change the estimates in the MIREC study. The increased precision using the average maternal U-F concentration as an indicator of prenatal fluoride exposure resulted in stronger statistical evidence of fluoride-associated deficits, as compared to the cross-sectional and the retrospective studies. Still, the exact fetal fluoride exposure during early brain development is not known, and even the maternal U-F concentrations may be considered somewhat imprecise. Such imprecision, likely occurring at random, will tend to underestimate fluoride neurotoxicity (Grandjean and Budtz-Jorgensen 2010).

The prospective studies offer strong evidence of prenatal neurotoxicity and should inspire a revision of water-fluoride regulations based on the benchmark results, especially for pregnant women and young children. Although systemic fluoride exposure may be associated with some benefits in dental health, these benefits appear to be small and non-essential prior to tooth eruption (Iheozor-Ejiofor et al. 2015), and other means of caries prevention, such as fluoridated toothpaste and other topical treatment, may be considered (Featherstone 2000).

## 5. CONCLUSIONS

Two prospective studies found concentration-dependent cognitive deficits associated with maternal U-F during pregnancy; one of the studies (Bashash et al. 2017) measured child IQ at two ages and found similar results, whereas the other study (R. Green et al. 2019) found a fluoride-IQ effect only in boys. We explored the shape of the concentration-response curve by using a standard linear shape and compared with a squared exposure and a piecewise linear function that allowed a change in steepness at two points within the range of exposures. The sensitivity calculations suggest that the standard linear function is a reasonable approximation. All of these estimates have a certain degree of uncertainty, and emphasis should therefore be placed on the joint BMC results from the two studies and involving both sexes. These findings, using a linear concentration dependence, suggest an overall BMCL for fluoride concentrations in urine of approximately 0.2 mg/L or slightly below that level.

## Data Availability

The availability of data has been stated in regard to the publications that our study relies on.

## ACKNOWLEDGMENTS

The ELEMENT study was supported by U.S. NIH R01ES021446, NIH R01-ES007821, NIEHS/EPA P01ES022844, NIEHS P42-ES05947, NIEHS Center Grant P30ES017885 and the National Institute of Public Health/Ministry of Health of Mexico. The MIREC study was supported by the Chemicals Management Plan at Health Canada, the Ontario Ministry of the Environment, and the Canadian Institutes for Health Research (grant # MOP-81285). PG is supported by the NIEHS Superfund Research Program (P42ES027706). CT is supported by the NIEHS (grants R21ES027044; R01ES030365-01).

The authors gratefully acknowledge: Nicole Lupien, Stéphanie Bastien, and Romy-Leigh McMaster and the MIREC Study Coordinating Staff for their administrative support, Dr. Jillian Ashley-Martin for providing feedback on the manuscript, as well as the MIREC study group of investigators and site investigators; Alain Leblanc from the INSPQ for measuring the urinary creatinine; Dr. Angeles Martinez-Mier, Christine Buckley, Dr. Frank Lippert and Prithvi Chandrappa for their analysis of urinary fluoride at the Indiana University School of Dentistry; Linda Farmus for her assistance with statistical modeling. This MIREC Biobank study was funded by a grant from the National Institute of Environmental Health Science (NIEHS) (grant #R21ES027044). The MIREC Study was supported by the Chemicals Management Plan at Health Canada, the Ontario Ministry of the Environment, and the Canadian Institutes for Health Research (grant # MOP-81285).

